# Radiologist-AI Collaboration for Ischemia Diagnosis in Small Bowel Obstruction: Multicentric Development and External Validation of a Multimodal Deep Learning Model

**DOI:** 10.1101/2025.09.05.25335014

**Authors:** Quentin Vanderbecq, Wen Fan Xia, Émilie Chouzenoux, Jean-Christophe Pesquet, Marc Zins, Mathilde Wagner

## Abstract

**Purpose:** To develop and externally validate a multimodal AI model for detecting ischaemia complicating small-bowel obstruction (SBO).

**Methods:** We combined 3D CT data with routine laboratory markers (C-reactive protein, neutrophil count) and, optionally, radiology report text. From two centers, 1,350 CT examinations were curated; 771 confirmed SBO scans were used for model development with patient-level splits. Ischemia labels were defined by surgical confirmation within 24 hours of imaging. Models (MViT, ResNet-101, DaViT) were trained as unimodal and multimodal variants. External testing was used for 66 independent cases from a third center. Two radiologists (attending, resident) read the test set with and without AI assistance. Performance was assessed using AUC, sensitivity, specificity, and 95% bootstrap confidence intervals; predictions included a confidence score.

**Results:** The image-plus-laboratory model performed best on external testing (AUC 0.69 [0.59–0.79], sensitivity 0.89 [0.76–1.00], and specificity 0.44 [0.35–0.54]). Adding report text improved internal validation but did not generalize externally; image+text and full multimodal variants did not exceed image+laboratory performance. Without AI, the attending outperformed the resident (AUC 0.745 [0.617–0.845] vs 0.706 [0.581–0.818]); with AI, both improved, attending 0.752 [0.637–0.853] and resident 0.752 [0.629–0.867], rising to 0.750 [0.631–0.839] and 0.773 [0.657–0.867] with confidence display; differences were not statistically significant.

**Conclusion:** A multimodal AI that combines CT images with routine laboratory markers outperforms single-modality approaches and boosts radiologist readers’ performance notably junior, supporting earlier, more consistent decisions within the first 24 hours.

**Key Points:** A multimodal artificial intelligence (AI) model that combines CT images with laboratory markers detected ischemia in small-bowel obstruction with AUC 0.69 (95% CI 0.59–0.79) and sensitivity 0.89 (0.76–1.00) on external testing, outperforming single-modality models.

Adding report text did not generalize across sites: the image+text model fell from AUC 0.82 (internal) to 0.53 (external), and adding text to image+biology left external AUC unchanged (0.69) with similar specificity (0.43–0.44).

With AI assistance both junior and senior readers improved; the junior’s AUC rose from 0.71 to 0.77, reaching senior-level performance.

**Summary Statement:** A multicentric AI model combining CT and routine laboratory data (CRP and neutrophilia) improved radiologists’ detection of ischemia in small-bowel obstruction. This tool supports earlier decision-making within the first 24 hours.

## INTRODUCTION

Acute small-bowel obstruction (ASBO) remains a high-stakes emergency in general surgery, accounting for 12–16 % of all surgical admissions and more than 300,000 operations each year in the United States alone (1–3). Direct hospital costs attributable to ASBO exceed US $2 billion annually, while emergency-general-surgery conditions as a group consume almost US $28 billion in national expenditure (4). Worldwide, the incidence of obstruction continues to rise, driven by postoperative adhesions, malignancy and an aging population (5).

Although most patients recover with nasogastric decompression or timely surgery, outcomes deteriorate precipitously once transmural ischemia develops: each hour of delay from hospital admission to operative intervention is associated with a 2–3 % absolute decrease in survival (6). Historical series report strangulation rates near 9 % and mortality approaching 8 % when ischemia is present (7), underscoring the urgent need to recognize ischemic bowel before irreversible necrosis or perforation occur.

Contrast-enhanced computed tomography (CT) is the gold standard imaging modality for suspected ASBO, offering unrivalled speed, availability and an ability to depict closed-loop configurations, mesenteric strangulation and indirect ischemia sign (8). A recent Chinese meta-analysis encompassing 45 studies and 4,004 patients reported that multidetector CT detects small-bowel ischaemia with pooled sensitivity 82 % (95 % CI 67–91 %) and specificity 92 % (86–95 %), with even higher sensitivity in adhesive SBO (96 % vs. 71 %, P = 0.05) (9). However, the diagnostic accuracy of individual CT signs remains inconsistent. Millet et al. found reduced mural enhancement to be highly specific (∼95 %) for strangulation, whereas other signs, like bowel wall thickening or mesenteric fluid, showed variable performance across studies(10). Some studies have also reported low diagnostic performance overall, like Sheedy et al. who found a sensitivity of only 15 %, increasing to 52 % after blinded consensus reinterpretation (11). Eze et al. identified a different pattern of significant CT predictors of ischemia in SBO, including peritoneal free fluid and bowel wall thickening, whereas Millet et al. emphasized decreased mural enhancement and mesenteric fluid(10,12). This discrepancy highlights the lack of consensus on key imaging features and supports the potential value of AI-assisted harmonization. Moreover, Eze’s findings suggest that integrating multimodal data, including clinical and laboratory variables, may further enhance diagnostic performance(12). Moreover, diagnostic confidence falls further during off-hour reporting (13). These real-world constraints create a diagnostic blind spot precisely where accuracy matters most.

Recent advances in artificial intelligence (AI) offer promising solutions. Three-dimensional vision model like convolutional neural networks (3D-CNNs) can interrogate entire CT volumes, learn complex perfusion patterns and output voxel-level risk maps within seconds. Early proof-of-concept studies have shown that deep-learning models outperform handcrafted machine-learning pipelines and approach expert accuracy for classifying obstruction severity (14), differentiating transient from irreversible ischemia in closed-loop obstruction (15), and even augmenting radiologist decision-making across diverse imaging tasks(16). Nevertheless, most prior work has been limited by small sample sizes and the absence of multimodal approach, leaving unanswered the critical question of whether AI can reliably identify ischemia before clinical deterioration.

In this multi-institutional retrospective cohort, we developed and externally validated a multimodal model that fuses volumetric CT features extracted by a 3D Vision transformer (ViT) with relevant laboratory markers. We hypothesize that this multimodal approach may improve diagnostic accuracy compared to current radiologic assessment alone, potentially enabling earlier and more accurate identification of ischemia.

## MATERIAL AND METHODS

This study was conducted and reported in accordance with the 2024 Checklist for Artificial Intelligence in Medical Imaging (CLAIM)(17). Institutional-review-board approval was obtained (IRB00012157 and IRB00011591).

### Datasets

#### Train and Validation Dataset

From the 10,240 abdominal CT examinations performed at our primary center between 2015 and 2022, 782 studies were excluded based on predefined criteria, and an additional 135 were excluded due to patient refusal of data use, resulting in 9,323 eligible scans. Exclusion criteria included non-exploitable images, incomplete abdominopelvic volumes, legal protection or guardianship status and patient objection to data use. An initial stratified random sample of 620 scans was selected for detailed annotation across study years. These labeled reports were then used to fine-tune a FlauBERT-based NLP model to identify likely bowel obstruction cases from free-text radiology reports.

To enrich the dataset for small bowel obstruction (SBO), we applied three parallel strategies:

1. Procedure-code review: all scans linked to a core set of surgical procedure codes (HGFA005, HGPC015, HGPA004, HPPC003, HPPA002) were included.
2. Diagnostic-code selection: ICD-10 codes associated with obstruction (K66, K31.5, K56.5, K91.3, K50, K43, R11, K55.9) were queried, and a subset was manually reviewed.
3. NLP filtering: the FlauBERT model classified report texts, from which a high-likelihood subset was manually validated.

From these strategies, a total of 1,171 scans were manually reviewed by a single observer with 4 years of post-attending experience in abdominal radiology (QV), and 77 additional cases with SBO Procedure codes from 2022–2025 were added, bringing the total to 1,248 CT scans. Among these, 669 were confirmed cases of small bowel obstruction.

To extend surgical SBO representation, we later added 102 additional cases from the second center, consisting of patients who had procedure-code of SBO between June 2022 and February 2025. Figure 1 display the flowchart of selection process.

**Figure 1:**
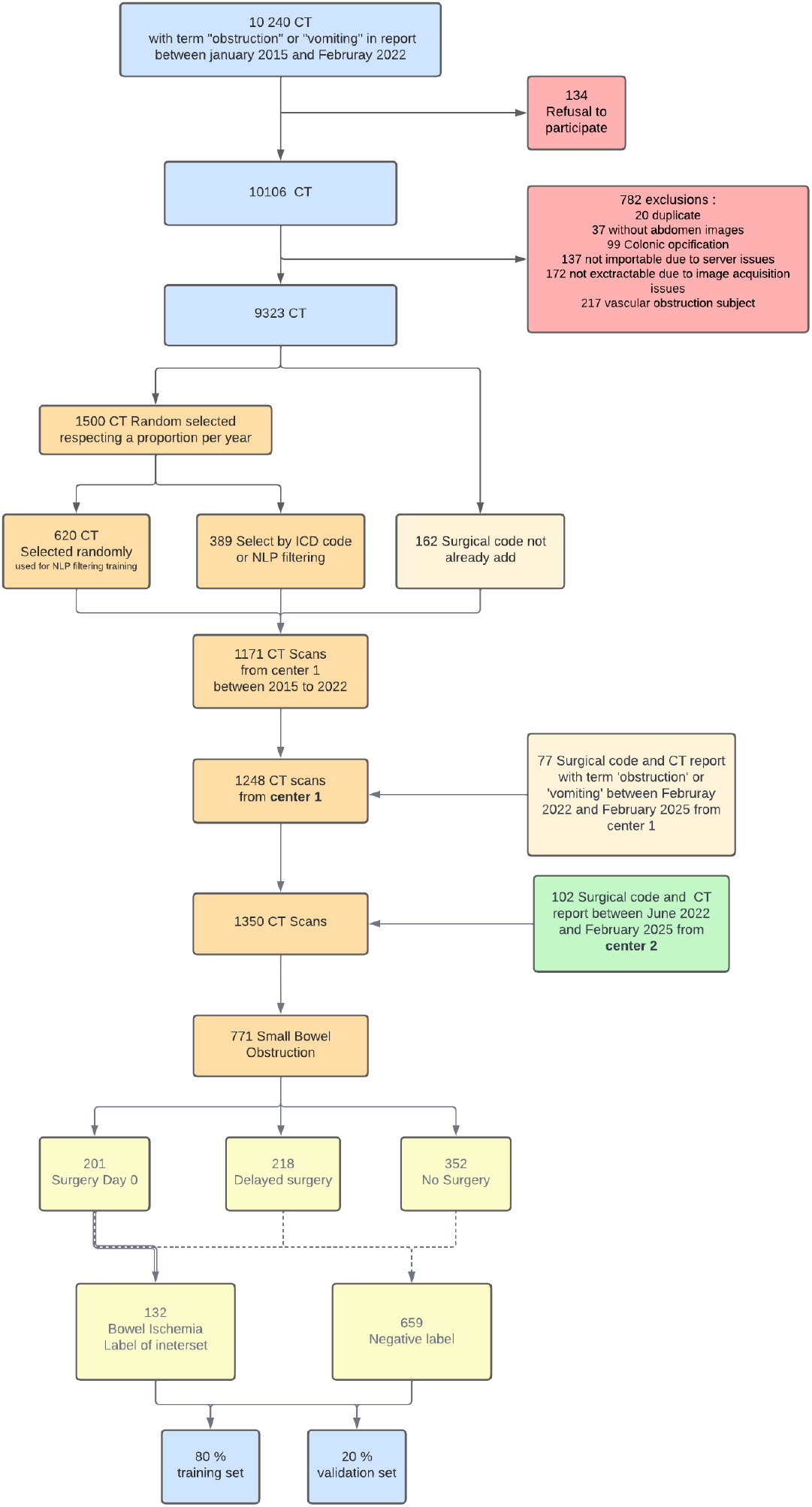
Flowchart. CT: Computed Tomography; ICD: International Classification of Diseases; NLP: Natural language processing

For model development we therefore used the 771 studies that demonstrated small-bowel obstruction. The remaining scans were split 80 %/20 % into training and validation sets with patient-level segregation. CTs were acquired on three GE Healthcare scanners (Revolution™ CT, Revolution™ EVO, Revolution™ Frontier) at the index institution and on Siemens Healthcare scanners at the two additional centers (Edge™ and Force™).

#### Objective judgment criteria for the classification task

Operative notes were reviewed for all patients who underwent surgery. Ischemia was labelled positive only when intra-operative ischemia was documented at surgery performed within 24 h of the index CT. The classification task was designed to predict the presence of this surgically confirmed ischemia.

#### External Validation Dataset

Patients from a third center were retrospectively screened between February 2022 and February 2025. Inclusion required either a surgical procedure for bowel obstruction or an admission coded ICD-10 K56. Of 80 eligible patients, 7 had no archived imaging, 2 were not usable due to technical transfer failure, and 5 were excluded owing to prior gastric-bypass surgery, these cases were omitted because the external validation center is a bariatric reference center, where this type of obstruction is overrepresented and rarely seen in the training or validation set. Moreover, gastric-bypass-related SBOs are considered clinically distinct, with ischemia present in nearly all cases (4 out of 5 in our cohort), which could introduce bias in performance assessment. Five gastric-bypass–related obstructions were excluded leaving 66 patients for external testing.

### Models overview

Three architectures processed images: a 3-D ResNet-101 whose pooling layer was replaced by an adaptive 4 × 4 × 5 kernel instead of 1 x 1 x 1, a multiscale vision transformer (MViT-3D) and a 3-D adaptation of the dual-attention ViT (18) (DaViT-3D). Each network was trained for 30 epochs with cross-entropy loss and stochastic-gradient descent; learning-rate, weight-decay and dropout were tuned by Bayesian search with Optuna (19) (supplementary table 1). All CT datasets were first limited to the abdomen and pelvis using a previously described pipeline (20). Voxel values were clipped to the 5th–95th percentile of each scan to dampen outliers, normalized (z-score), and resampled to isotropic cubes: 224×224×224 voxels for convolutional networks and 112×112×112 for transformer backbones. During training every volume had a 50 % chance of horizontal or vertical flipping and an independent chance of being rotated by 90°, 180° or 270°.

For the language branch, we retained only the clinical context paragraph—everything from “Indication” (or “History”) up to “Technique”, or to “Findings” if the technique header was absent. The text was lower-cased, accents and other diacritics were stripped, then tokenised with FlauBERT-base, a French BERT model whose pretrained weights served as the initial state for fine-tuning.

Two routinely ordered blood tests, C-reactive protein and absolute neutrophil count, were used in the biological branch. When either value was missing, we inserted a fixed placeholder (0 mg L^-1^ for CRP, 10 G L^-1^ for neutrophils) and appended a binary flag so the network could learn to distinguish real from imputed measurements.

Image, text, and biology features were each projected into a 512-dimensional latent space. Whenever more than one source was present, their embeddings were summed, subjected to a tanh activation and dropout, and fed to a single linear classifier. Alongside the probability of ischemia, the system emitted a confidence score derived from the distance between that probability and the 0.5 decision threshold; predictions closer to the boundary were tagged as less certain.

Further architectural details (Stext), optimization ranges (Stable 1) and the final hyper-parameter configurations (Stable 2) are provided in the Supplementary Material.

### Radiologists’ image analysis

To compare the performance of the model and the one of the radiologists, different analyses were done. For real-world assessment, radiology reports were analyzed by a single radiologist, with 13 years of experience in abdominal radiology, and the mention-signs of ischemia were recorded.

The performance of a senior and a junior radiologist were also tested. The senior radiologists were an attending radiologist with 4 years of post-residency experience in abdominal radiology, and the junior was a resident, who each read independently the cases of the external testing cohort. They were asked to identify the presence of bowel ischemia. Each case was read with three successive passes: first without assistance, then with access to the model’s prediction, and finally with both the model’s prediction and its associated confidence score.

### Statistical analysis

To evaluate model performance, we computed balanced accuracy, calculated as the mean of sensitivity and specificity, and the F1 score defined as 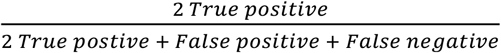or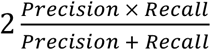, and the area under the ROC curve (AUC-ROC) computed from the final model score. Given the class imbalance in the dataset, balanced accuracy was used as the primary performance metric on validation set, particularly for hyperparameter optimization with Optuna(19). Model computations were performed using the PyTorch framework for training and validation sets, while test set evaluations utilized the scikit-learn Python library. We estimated 95 % confidence intervals for both model and radiologist performance using 1000 bootstrap resamples.

## RESULTS

### Dataset

Training-validation set included 771 cases of small bowel occlusion, 687 were mechanical and 84 functional. The cohort included 409 women (53%); the mean age of the overall cohort was 67 years. Among the occlusions 711 patients, 186 (26%) patients underwent surgery on the day of diagnosis (Day0), 190 (27%) had surgery delayed by over 24 hours following CT imaging (Delayed surgery), while 335 (47%) were managed conservatively. Of those operated on Day0, 130 (18%) experienced documented ischemia and 67 (9%) underwent bowel resection due to irreversible ischemia and necrosis. Some details of etiology are described in Table 1.

**Table 1:**
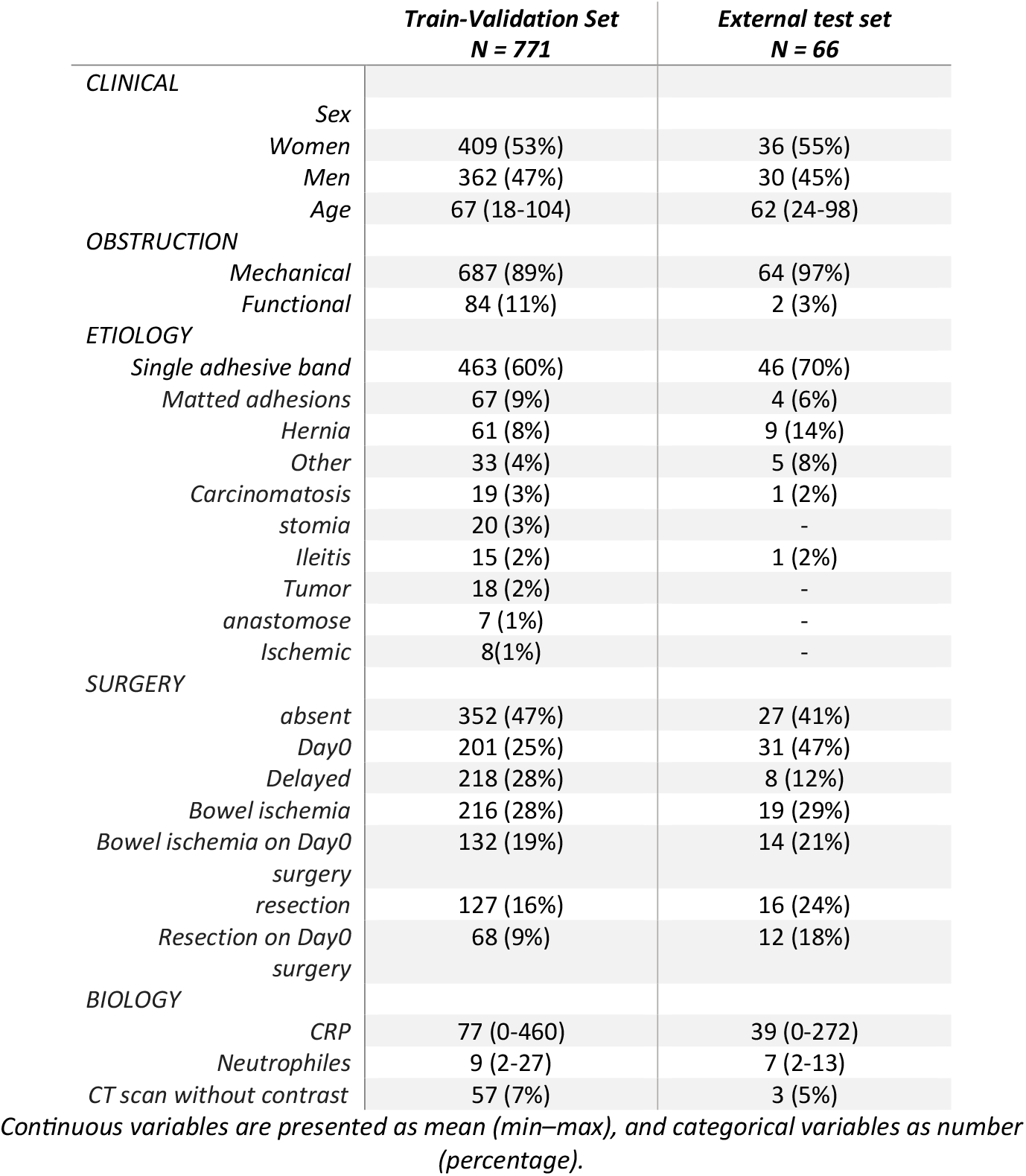
Characteristics of the patients.

In the external test set of 66 cases, all were small bowel obstructions, 65 mechanical and 1 functional. The external test set included 36 women (55%); the mean age of this external set was 62 years. Among the SBO, 31(47%) patients underwent surgery on the day of diagnosis (Day0), and 14 (21%) showed signs of ischemia. Eight patients had delayed surgery, while the remaining 27(41%) were managed conservatively.

### Models’ performances

The three image-only backbones, MViT-3D, DaViT-3D and ResNet-101-3D, were benchmarked on the validation and external test sets. MViT-3D showed the best validation performance (AUC 0.68, sensitivity 0.77) and was therefore selected as the reference image model.

On external testing, MViT-3D achieved AUC 0.53 [0.41–0.64] with sensitivity 0.76 [0.61–0.88] and specificity 0.37 [0.27–0.47]. DaViT-3D and ResNet-101-3D performed comparably (AUC 0.50 [0.40–0.61] and 0.52 [0.40–0.63], respectively). A biology-only multilayer perceptron reached AUC 0.63 on internal validation but fell to 0.51 [0.34–0.67] on the test set. The text-only model, despite strong internal metrics (AUC 0.90), generalized poorly (test AUC 0.52 [0.35–0.68]) (Table 2).

**Table 2:** Models performances.

|  | VALIDATION SET |  |  | EXTERNAL SET |  |  |
| --- | --- | --- | --- | --- | --- | --- |
|  | AUC | Specificity | Sensitivity | AUC | Specificity | Sensitivity |
| **IMAGE MODEL** |  |  |  |  |  |  |
| - MVIT | 0.68 | 0.59 | 0.77 | **0.53 [0.41-0.64]** | 0.37 [0.27-0.47] | 0.76 [0.61-0.88] |
| - DAVIT | 0.65 | 0.58 | 0.72 | **0.50 [0.40-0.61]** | 0.45 [0.36-0.55] | 0.50 [0.33-0.66] |
| - RESNET101 | 0.63 | 0.57 | 0.69 | **0.52 [0.40-0.63]** | 0.53 [0.44-0.63] | 0.56 [0.38-0.74] |
| **BIOLOGY MODEL** | 0.63 | 0.88 | 0.38 | **0.51 [0.34-0.67]** | 0.15 [0.06-0.25] | 1 [1-1] |
| **TEXT MODEL** | 0.90 | 0.93 | 0.87 | **0.52 [0.35-0.68]** | 0.56 [0.42-0.70] | 0.57 [0.30-0.82] |
| **MULTIMODAL** |  |  |  |  |  |  |
| - MVIT/TEXT/BIOLOGY | 0.66 | 0.55 | 0.77 | **0.69 [0.59-0.79]** | 0.43 [0.34-0.53] | 0.89 [0.76-1.0] |
| - MVIT /BIOLOGY | 0.67 | 0.65 | 0.69 | **0.69 [0.59-0.79]** | 0.44 [0.35-0.54] | 0.89 [0.76-1.0] |
| - MVIT /TEXT | 0.82 | 0.79 | 0.85 | **0.53 [0.41-0.64]** | 0.37 [0.27-0.47] | 0.76 [0.61-0.88] |
| - TEXT/BIOLOGY | 0.91 | 0.90 | 0.92 | **0.49 [0.34-0.65]** | 0.32 [0.19-0.44] | 0.80 [0.6-1.0] |

The values in brackets represent the 95% confidence intervals, calculated using the bootstrap method with 1,000 samples. MVIT: Multiscale vision transformer. DaViT: dual attention vision transformer. Text model was Flaubert base model that was fine tune. Biology model was multi-layer perceptron trained on CRP and neutrophils levels.

Combining modalities improved performance. The image + biology configuration yielded the highest external AUC at 0.69 [0.59–0.79] with sensitivity 0.89 [0.76–1.00] and specificity 0.44 [0.35–0.54]. Adding text to this pair (image + text + biology) did not change AUC (0.69 [0.59– 0.79]) but left specificity similar (0.43 [0.34–0.53]). The image + text variant retained good internal AUC (0.82) yet tested at 0.53 [0.41–0.64] (Table 2; Figure 2 and 3).

**Figure 2:**
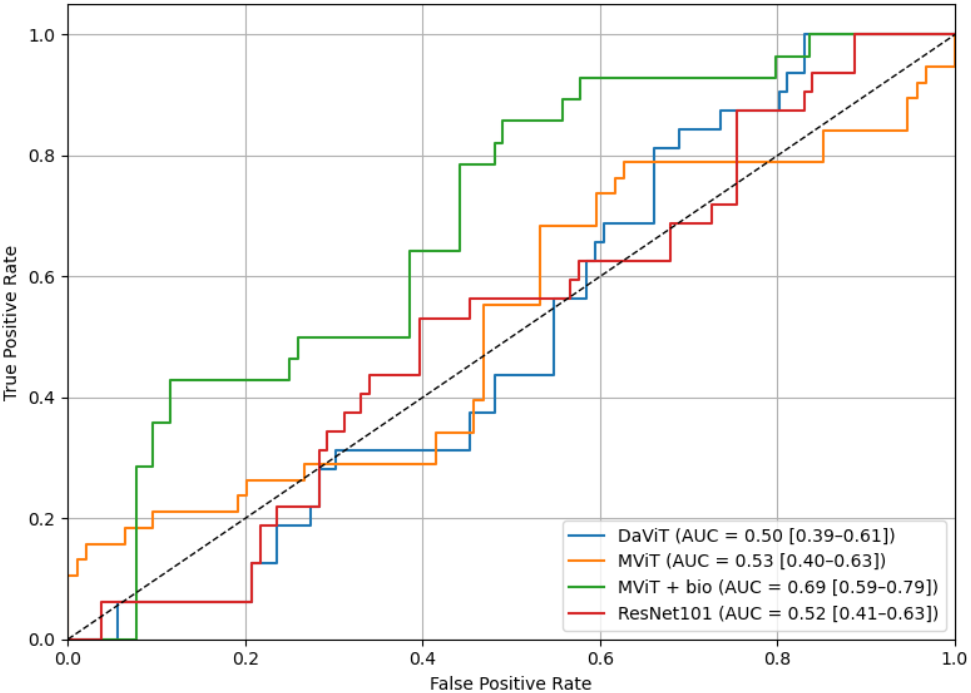
ROC Curve of image model and MViT/biology model. AUC: Area Under the Receiver Operating Characteristic Curve. 95% confidence interval in brackets. MViT: Multiscale vision transformer. DaViT: dual attention vision transformer.

**Figure 3:**
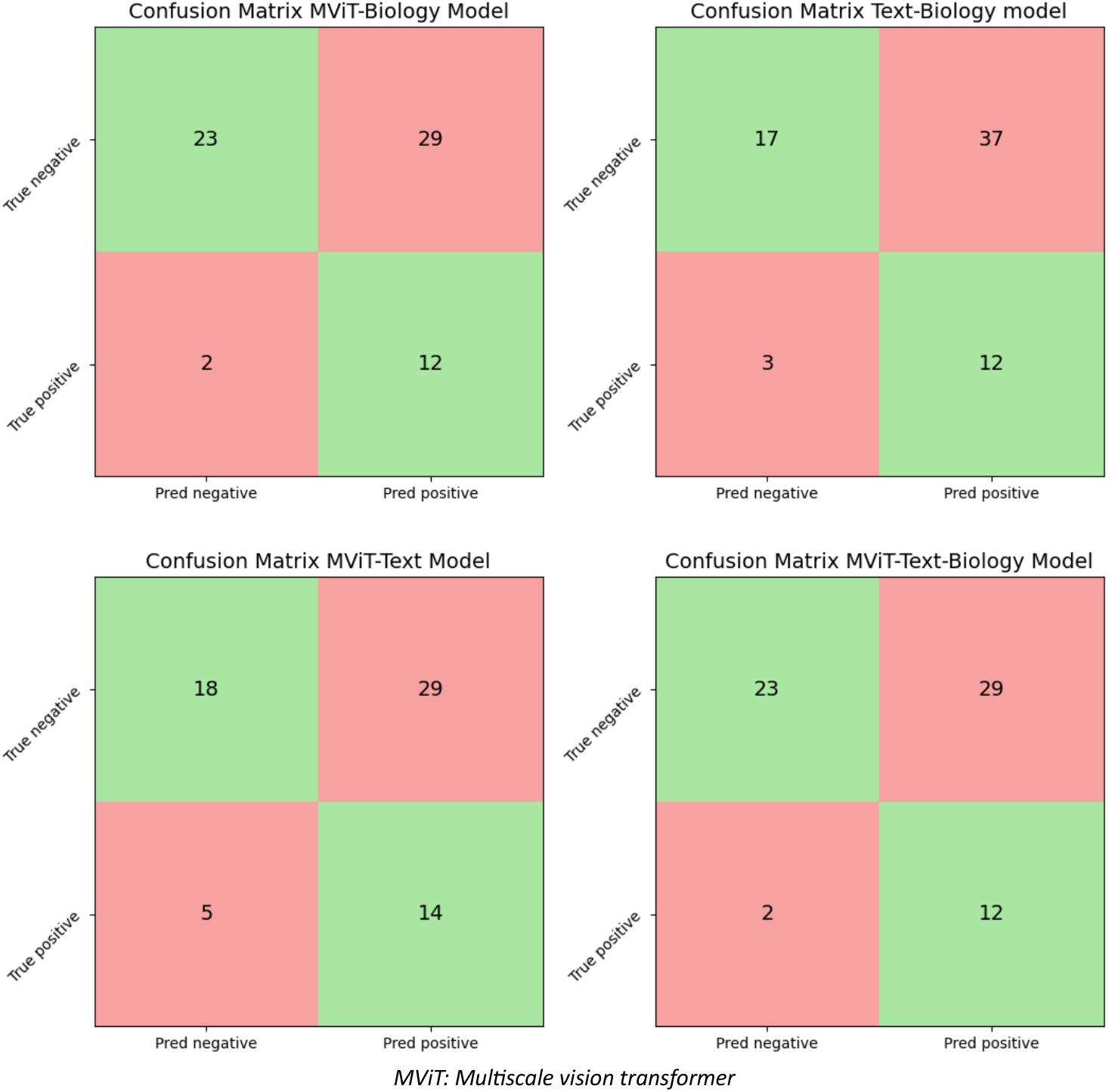
Confusion matrix of multimodal models.

### Evaluation of the radiologists and the radiologist augmented’s performance

Results are presented in Table 3. In the real-world setting from report description the AUC for identifying bowel ischemia was 0.736 (95 % IC [0.612–0.847]), with a sensitivity of 0.722 (95 % IC [0.500–0.923]) and a specificity of 0.748 (95 % IC [0.648–0.842]). The senior radiologist’s performance was slightly higher, with an AUC of 0.745 (95 % IC [0.617–0.845]), sensitivity of 0.714 (95 % IC [0.500–0.917]), and specificity of 0.769 (95 % IC [0.673–0.854]) while the one of the resident was slightly lower with an AUC of 0.706 (95 % IC [0.581–0.818]), sensitivity of 0.727 (95 % IC [0.500–0.909]) and specificity 0.686 (95 % IC [0.585–0.792])(Table 3).

**Table 3:**
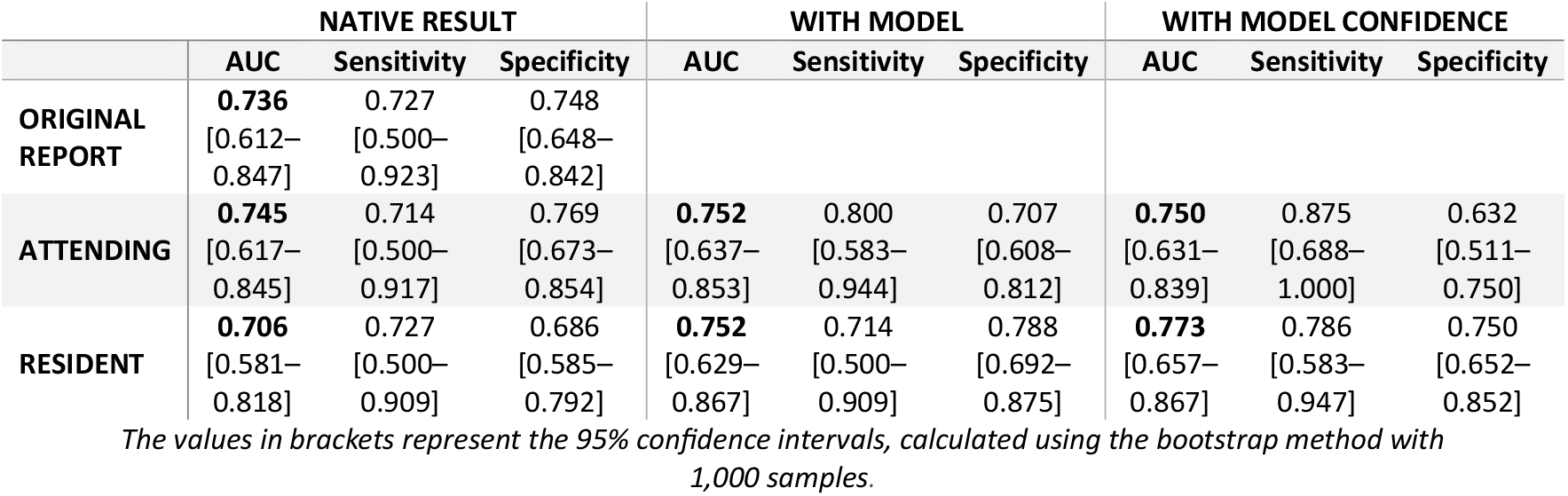
Radiologists and augmented radiologists’ performance.

Model assistance improved both readers’ performance. For the attending, AUC increased to 0.752 [0.637–0.853] with the model, and to 0.750 [0.631–0.839] when confidence scores were shown, with sensitivity rising from 0.714 to 0.875 but at the cost of decreased specificity. For the junior, AUC rose to 0.752 [0.629–0.867] with model support, matching the attending’s performance, and further improved to 0.773 [0.657–0.867] with confidence display, alongside gains in both sensitivity and specificity. Although no statistically significant difference was observed between conditions based on overlapping 95% confidence intervals from bootstrap estimates (Table 3).

## DISCUSSION

We have developed a multimodal AI model designed to improve diagnostic accuracy for ischemia in small bowel obstruction (SBO). This model integrates imaging data with laboratory inputs to support radiologists in making more confident and precise diagnoses. Beyond diagnostic accuracy improvement, the AI tool fosters improved communication between radiologists, surgeons, and clinicians, helping to guide therapeutic decisions and increase confidence in treatment strategies.

Recent advances in medical machine learning have highlighted the value of multimodal data integration, combining imaging with clinical and biological data, in order to improve diagnostic accuracy beyond unimodal approaches(21,22). In clinical practice, physicians already synthesize imaging with lab results to refine diagnoses, and AI models increasingly emulate this strategy. However, for specific conditions such as mechanical bowel obstruction, few have attempted to use multimodal data, perhaps because abdominal CT features have proven more predictive of clinical outcomes than any single biological marker. For example, a 2017 study showed that CT findings and radiologist assessments outperformed clinical signs and blood markers like lactate in predicting urgent surgical need.(23)

Internal validation showed the language-only model to be highly accurate, but its performance dropped sharply on the external test set and it provided no added value to the multimodal network. This lack of generalization likely results from three interconnected factors. The first concerns domain-specific language: the vast majority (92%) of training notes came from a single hospital, where clinicians are likely to use a consistent and highly localized set of abbreviations and phrasing. Such linguistic uniformity can hinder a model’s ability to adapt to other institutions with different documentation styles. The second issue is limited data volume by deep learning standards. Despite eight years of bowel obstruction cases from the primary hospital (handling around 75,000 emergency visits annually) and three years from two additional centers with over 65,000 visits each, the number of positive cases remains far below the scale typically required to train robust Transformer-based models, for example, the CheXpert chest-X-ray corpus alone contains 224 000 studies from 65 000 patients(24). The third factor involves imaging data scarcity. All vision models achieved strong balanced accuracy on the training set, but consistently overfit and performed poorly on internal validation scans, with further degradation on external data.(25) This situation is further complicated by the sensitive nature of medical data, whereby there is imposed strict privacy, regulatory, and ethical constraints. This makes it difficult to pool large-scale datasets across institutions. Sharing imaging and clinical data across hospitals often requires complex data governance agreements, pseudonymization protocols, and infrastructure capable of hosting and processing high-resolution 3D scans(26). Moreover, training advanced multimodal models, especially those incorporating full 3D CT volumes, demands substantial computational resources, including high-memory GPUs and centralized servers, which are not uniformly available across hospitals. Without coordinated multi-institutional efforts and dedicated infrastructure, expanding datasets to the scale needed for generalizable AI remains a major challenge.

This study has several limitations. First of all, the decision to include AN and CRP as biological markers is open to debate. Several studies highlight the prognostic value of the total white-blood-cell count (WBC) without separately analyzing the neutrophil fraction, even though WBC and neutrophils are closely correlated (12,27,28). Because one study reported an even stronger association for neutrophils, we opted to retain this variable (29). CRP emerges less consistently in the literature, it does not reach statistical significance, yet, after procalcitonin (PCT), which is seldom measured routinely in French hospitals, CRP remains the most informative and best-standardized inflammatory protein. Secondly, our ischemia label is probably biased and not ideal, for several reasons. First, the label rests on surgical confirmation within 24 hours of the index CT, even though ischemia may develop after the CT examination. Second, operative notes lack a standardized vocabulary for small-bowel ischemia, so some truly positive cases are probably recorded as negative. Third, some patients who showed early radiological or clinical signs of bowel compromise were not taken to surgery during the first 24 hours and were therefore assigned to the negative cohort. These two latter scenarios are the most plausible and would introduce false negatives, suggesting that our model, with high sensitivity, might actually perform better than the current metrics imply if real-time monitoring of small-bowel viability were available.

To conclude, a multimodal AI tool that fuses CT with routine laboratory data improves ischemia detection in SBO and helps reduce reader variability. In the emergency setting, where confirming ischemia within the first 24 hours is critical, AI assistance can support earlier, more reliable triage and decision-making. Future work should prioritize generalizable text integration and prospective, multicenter validation.

## Supporting information

Stext

## Data Availability

All data produced in the present study are available upon reasonable request to the authors

