## Supplementary material for "Radiologist-AI Collaboration for Ischemia Diagnosis in Small Bowel Obstruction: Multicentric Development and External Validation of a Multimodal Deep Learning Model": Stext

***Supplementary text: Models and training procedure description***

ResNet-101-3D retained the original 2 048 channels but replaced global average pooling with an adaptive 4 × 4 × 5 kernel, producing a 2 048 × 4 × 4 × 5 tensor that was flattened before the final fully connected layer. MViT-3D (multiscale vision transformer 3D) was ported directly from the official PyTorch video implementation with little adjustment to could process a high third dimension (112), whereas DaViT-3D (Dual attention vision transformer) followed Ding et al. readapt to 3D images.

All models were trained for 30 epochs with cross-entropy loss and stochastic-gradient descent. We used a triangular CycleLR schedule (base $1 \times 10^{-5}$, peak $1 \times 10^{-3}$, step-up 15) in your hyper-parameter search. Previously we evaluate others scheduler as Cosine, reduce on plateau, with hight performance with CycleLR on MViT and Resnet models and decide to only used it in further experience. During training each volume had a 50 % chance of horizontal or vertical flip and an independent chance of being rotated by 90°, 180° or 270°. Random gaussian noise and texture get provide in second step of training

Optuna carried out two sequential 20-trial searches: the first minimized validation loss, the second maximized balanced accuracy. Learning rate was explored from $1 \times 10^{-4}$ to $1 \times 10^{-2}$; weight decay from $1 \times 10^{-4}$ to $1 \times 10^{-1}$ (log-uniform); momentum from 0.50 to 0.99; and dropout from 0.10 to 0.80 in convolutional blocks or 0.60–0.90 in fusion heads. For text fine-tuning, dropout ranged 0.10–0.80 in the initial run and 0.30–0.80 after data augmentation (synonym substitution, random deletion and word swapping), in second.

The biology multilayer perceptron comprised two hidden layers with ReLU activations, batch normalization and dropout; hidden sizes were searched between 32–256 and 16–128 units. All modality-specific embeddings were projected to 512 dimensions using linear layers initialized with Xavier-uniform weights.

Prediction confidence was calibrated with a Gaussian mapping: probabilities were treated as z-scores centered on the 0.5 decision threshold so that ± 2 σ contained 95 % of outputs, values near 0.5 thus received low confidence, and those nearer 0 or 1 high confidence.

Models were trained and validated on an NVIDIA RTX A6000 GPU (48 GB).

**Supplementary table 1: Hyper-parameter Search Ranges**

| *Model / Variant* | *Learning-rate* | *Weight-decay* | *Dropout (image / fusion / text)* | *Other tunables* |
|---|---|---|---|---|
| ***ResNet-101-3D*** | $1 \times 10^{-4} – 1 \times 10^{-2}$ | $1 \times 10^{-4} – 1 \times 10^{-2}$ | 0.10 – 0.80 / — / — | Momentum 0.50 – 0.99; scheduler on/off |
| *MViT-3D* | $1 \times 10^{-4} – 1 \times 10^{-2}$ | $1 \times 10^{-4} – 1 \times 10^{-2}$ | 0.10 – 0.80 / — / — | Momentum 0.50 – 0.99; scheduler on/off |
| *DaViT-3D* | $1 \times 10^{-4} – 1 \times 10^{-2}$ | $1 \times 10^{-4} – 1 \times 10^{-2}$ | 0.10 – 0.80 / — / — | Momentum 0.50 – 0.99; scheduler on/off |
| *Biology MLP* | $1 \times 10^{-4} – 1 \times 10^{-2}$ | $1 \times 10^{-4} – 1 \times 10^{-2}$ | 0.10 – 0.80 | Hidden-1: 32–256, Hidden-2: 16–128; scheduler on/off |
| *Text (FlauBERT)* | $1 \times 10^{-5} – 1 \times 10^{-3}$ | $1 \times 10^{-4} – 5 \times 10^{-2}$ | — / — / 0.10 – 0.80 | Scheduler on/off; data-augmentation on/off |
| *Image + Biology* | $1 \times 10^{-4} – 1 \times 10^{-2}$ | $1 \times 10^{-4} – 1 \times 10^{-1}$ (log scale) | 0.60 – 0.90 / 0.60 – 0.90 / — | Momentum 0.50 – 0.99; scheduler on/off |
| *Image + Text* | $1 \times 10^{-4} – 1 \times 10^{-2}$ | $1 \times 10^{-4} – 1 \times 10^{-1}$ | 0.10 – 0.80 / 0.60 – 0.90 / 0.60 – 0.90 | Momentum 0.50 – 0.99; scheduler on/off |
| *Image + Text + Biology* | $1 \times 10^{-4} – 1 \times 10^{-2}$ | $1 \times 10^{-4} – 1 \times 10^{-1}$ | 0.60 – 0.90 / 0.60 – 0.90 / 0.60 – 0.90 | Momentum 0.50 – 0.99; scheduler on/off |
| *Text + Biology* | $1 \times 10^{-4} – 1 \times 10^{-2}$ | $1 \times 10^{-4} – 1 \times 10^{-1}$ | — / 0.60 – 0.90 / 0.60 – 0.90 | Text-projection 128–512; scheduler on/off |

Scheduler: CycleLR (base_lr=1e-5, max_lr=1e-3, step_size_up=15, mode="triangular").

**Supplementary table 1: Best Hyper-parameters**

| *Model (backbone)* | *LR* | *Weight decay* | *Dropout (image / fusion / text)* | *Momentum / Scheduler* | *Hidden units / Extras* |
|---|---|---|---|---|---|
| ***ResNet-101-3D*** | $2.0 \times 10^{-3}$ | $1.2 \times 10^{-3}$ | 0.30 / — / — | 0.90, CycleLR (base $1 \times 10^{-5}$ → max $1 \times 10^{-3}$) | — |
| ***MViT-3D*** | $1.5 \times 10^{-3}$ | $4.0 \times 10^{-4}$ | 0.30 / — / — | 0.90, none | — |
| ***DaViT-3D*** | $1.8 \times 10^{-3}$ | $8.0 \times 10^{-4}$ | 0.35 / — / — | 0.90, none | — |
| ***Biology MLP*** | $1.3 \times 10^{-3}$ | $1.0 \times 10^{-3}$ | 0.15 | — | Hidden layers 168 → 56 |
| ***Text (FlauBERT)*** | $1 \times 10^{-5}$ | $1.0 \times 10^{-2}$ | — / — / 0.34 | — / scheduler on | — |
| ***Image + Biology*** | $2.66 \times 10^{-3}$ | $8.83 \times 10^{-2}$ | 0.74 / 0.76 / — | 0.81, scheduler on | — |
| ***Image + Text*** | $8.91 \times 10^{-3}$ | $1.10 \times 10^{-3}$ | 0.58 / 0.90 / 0.61 | 0.81, scheduler on | — |
| ***Image + Text + Biology*** | $9.8 \times 10^{-3}$ | $1.35 \times 10^{-2}$ | 0.73 / 0.90 / 0.61 | 0.81, scheduler on | — |
| ***Text + Biology*** | $2.45 \times 10^{-3}$ | $3.25 \times 10^{-3}$ | — / 0.73 / 0.69 | — / scheduler off | Text projection 384 |
